# Clinical and sociodemographic predictors of hearing aid use in infants aged 0-2 with permanent childhood hearing loss: a retrospective cohort single-site pilot study

**DOI:** 10.1101/2023.05.31.23290669

**Authors:** Emilee Gosnell, Saima Rajasingam, Merle Mahon, Deborah Vickers

## Abstract

**Objectives:** The aim of this study was to identify differences in hearing aid use among infants aged 0-2 years with respect to clinical and sociodemographic factors in an attempt to better predict which patients and families may benefit from extra support in the early stages of hearing aid fitting to encourage optimum usage. The secondary aim was to investigate how hearing aid use changed over the first two years post-fitting.

**Methods:** A retrospective review of records was performed on 252 patients, aged 0-2 years with permanent childhood hearing loss from a single-site hospital who were fitted with hearing aids between 2005-2022. Ninety-six patients met the inclusion criteria. Datalogging values were collected for six different time points post fitting that coincided with their routine clinical follow-up appointments: 2 weeks, 6 weeks, 6 months, 12 months, 18 months and 24 months. Clinical and sociodemographic information was also collected for each participant. This included sex, average pure tone threshold, unilateral vs bilateral use, speech intelligibility score, additional disabilities, Index of Multiple Deprivation, Income Decile, Education and Skills Decile, Income Deprivation Affecting Children Index (IDACI), ethnicity and home language.

**Results:** The datalogging results indicated a median average of 4.67 hours (3.0-7.3) hours per day use across patients and across the first two years post-fitting. Differences in datalogging according to IDACI Decile was significant (p=0.01), suggesting that infants from the more deprived groups (1-5) used their devices less. All other predictors did not reach statistical significance. There was insufficient data to investigate change in hearing aid use over the first two years post-fitting.

**Discussion:** There were significant amounts of missing datalogging information. Some of the missing data was attributed by the clinical teams to lost hearing aids or unattended appointments. It is also believed that time restraints in clinic are the primary barrier. The datalogging values are lower than expected. It is considered that up to 12 hours per day use is necessary for good speech and language development. The findings highlighted that many families struggle to achieve optimal hearing aid usage in their infants. There was an association between daily device use and one socioeconomic status predictor.

**Conclusions:** Average datalogging values for daily hearing device use were lower than considered optimal for 0-2 year olds with permanent childhood deafness. The only factor related to these findings was IDACI Decile indicating that infants from more deprived backgrounds may achieve lower hearing aid usage than those from the lesser deprived regions. This finding needs to be verified on a larger scale and better understood to explore potential approaches to overcome the problems. The role of multiple factors also needs to be explored with a larger sample size. This would require a multi-centre study to be conducted.

## Introduction

The UK National Hearing Screen Program (NHSP) was fully implemented in 2006 with a vision to improve outcomes through widespread hearing screening, effective diagnostic assessments and family-centred early intervention [1]. The mean age of hearing aid fitting decreased substantially from 26.3 months [2] in the period 1985-1993 to 82 days for the 2012/2013 UK birth cohort [1].

Early intervention and access to aiding is crucial for infants with hearing loss to enable speech and language acquisition within the critical period, particularly for those with increasingly higher hearing thresholds [3]. Further impacts on quality of life, socio-emotional development and early learning have also been highlighted [4].

Despite advances in the early intervention pathway, the benefits of aiding may only be realised with consistent daily usage [5]. Full-time use of hearing aids has been defined as more than 12 hours per day [6]. It is accepted that infants’ sleeping patterns can affect this number, particularly in the newborn stage where hearing aids are now often issued, and infants may only be alert for 1-2 hours at a time. A few studies have investigated duration of hearing aid use with subjective parental estimation or objective datalogging measures and found wide variation in the pattern of results, also with differing definitions of consistent usage [6-12]. Two studies by Muñoz et al. [9,10] indicated 38% and 43% of parents reporting use of aiding as less than all waking hours in children aged 0 to 18 months and 19 to 36 months respectively. Further, interviews with 7 families performed by Moeller et al. [8] found that only 3 families were able to achieve consistent use of hearing aids by the age of 16.5 months. Fitzpatrick et al. [7] observed in a retrospective chart review that consistent hearing aid use was achieved in the home environment by 51% of children with unilateral, mild and high frequency bilateral hearing loss in their sample of 5000, although age was not specified. There was further evidence that 36.7% of children showed inconsistent or no usage. In contrast, a study performed in Australia with a sample size of 413 children with hearing aid and cochlear implants found that according to parental reporting, 71% of children wore their devices for over 8 hours per day at 3 years of age [11].

The issues regarding parental report of hearing aid usage have been discussed. Walker at al. [12] compared parental reports of use with hearing aid datalogging and found that 84% of parents overestimated use by 2.6 hours per day (SD=2.52) and 16% underestimated use.

Datalogging capabilities are available in most current hearing aid models whereby the instrument records the average number of hours per day a hearing aid is on. The information can be extracted in the clinic from the hearing aid software and is often used as an assessment and counselling tool to gauge family engagement with the user’s rehabilitation plan. Datalogging averages use by the number of total hours switched on and days between last and current connection to the software. Therefore, periods where the hearing aid(s) may not have been worn, for example due to a fault, will be taken into account and reduce the daily use average. Similarly, datalogging can overestimate the child’s use if the hearing aid is not switched off when it is removed. When using datalogging values in the research design, two studies have highlighted daily usage of the hearing aid at 4.36 hours per day (SD=3.17) for infants aged 6-24 months [12] and infants aged 0-4 as 5 hours per day [6]. In both studies, hearing aid use increased for children of school age.

The impact of family socioeconomic status on hearing aid usage has been explored. Three studies found that poorer hearing, older age and higher maternal education level was associated with higher hearing aid usage in children [5,11-12]. In addition, Marnane et al. identified an association of higher socioeconomic status measured by the Index of Relative Socioeconomic Advantage and Disadvantage (IRSAD), earlier age of intervention and higher number of early intervention hours with higher device usage in the hearing aid sub-group [11]. Other studies have attempted to explore socioeconomic predictors of hearing aid usage in the US, with health insurance status being the critical variable. Small sample sizes, difficulties in accessing data and participant recruitment from mostly white, educated and higher income families also limit this. Much of the literature which explores parent-reported challenges to optimal hearing aid use focuses on situational and practical aspects of daily use [13,14] but often fails to capture cultural, linguistic and social barriers. A study which looked at hearing aid usage and challenges among children from Hispanic families, with many parents of lower income and education status within their participant group, discovered that 66% wore their hearing aids all day on ‘good days’. Thirty-seven per cent were wearing their hearing aids all day on ‘bad days’ [15]. The investigators commented that datalogging information would be helpful but was not included.

It is well accepted that the prevalence of childhood hearing loss is higher in ethnic minority and socioeconomically disadvantaged communities. This may be due to a higher incidence of prematurity and low birth weight; economic, educational and employment disadvantages associated with parents who have long-standing hearing loss, incorporating genetic and familial factors as a cofounder; and in many South Asian and Middle Eastern migrant communities where consanguineous marriage is more prevalent, thus increasing the risk of homozygous traits in their offspring [16-19].

The aims of this study are:

1. To quantify average hearing aid use in infants aged 0-2 within our department’s patient cohort through collection of datalogging values.
2. To investigate whether hearing aid use time increases within the first two years in infants fitted aged 0-2.
3. To assess the feasibility of collecting datalogging, and clinical and sociodemographic information to explore associations with hearing aid use.
4. To collect data which can inform a sample size calculation for a larger multicentre study with the aim of highlighting patients and families which are at higher risk of low hearing aid usage.

## Materials and methods

Data used in the present study was collected from a case records review. Data was collected in January 2023.

### Ethical considerations

All approvals were granted by the Health Research Authority (REC reference: 22/HRA/2309).

### Participants

Patients with permanent childhood hearing loss were identified using the Auditbase (Auditdata, Denmark) patient management software at the Audiology Department of St George’s University Hospital NHS Foundation Trust in Southwest London, UK. It is a large inner city hospital.

Data for 252 patients with permanent hearing loss cared for at the department between 2005-2022 was extracted. Patients were included in the study if they were fitted with hearing aids before the age of 2 years and if they wore conventional air-conduction hearing aid(s) with datalogging capabilities and available data. Of the 252 identified patients, 158 were excluded from the study. Reasons for exclusion included: a diagnosis of ANSD where hearing aids were not indicated (n=4); a bone conduction or bone anchored device was prescribed (n=21); patients were fitted post-2 years of age (n=30); cases where there was no datalogging information available (n=13); a late-identified or late-onset hearing loss post-2 years of age (n=37); patients who transferred in after or out of the service before their initial hearing aid fitting (n=31); a diagnosis of mild or unilateral hearing loss where hearing aids were not fitted (n=19); a patient who went private (n=1) and two cases where the patient sadly died. A total of 94 patients were included in the study. All data was pseudonymised on the raw data sheet.

### Study outcomes

The primary outcome for this study was hearing aid usage as quantified by hearing aid datalogging.

### Clinical information

Information at the initial fitting was collected from Auditbase: patient age in months, pure-tone average thresholds, speech-intelligibility index score (SII) and whether the fitting was unilateral or bilateral. Pure-tone average threshold was calculated from frequencies 0.5, 1, 2 and 4kHz. In some cases, a pure-tone threshold at 2kHz was not available (n=13) and averaging was performed using the three remaining thresholds.

Presence of additional disabilities of the child including global developmental delay, autism, ADHD, syndromic conditions and physical disability were recorded. Some of the additional disabilities were not known at the time of initial fitting and were noted during retrospective review of the medical records at the point of data collection. This is important to bear in mind as the absolute number of additional disabilities among participants may increase after this manuscript goes to press.

Datalogging was extracted from the patient’s written records or hearing aid software across six different time points post-fitting: 2 weeks, 6 weeks, 6 months, 12 months, 18 months and 24 months. These time points were specified and controlled for during the study according to the routine clinical follow-up procedure for the child. Due to common issues with appointment scheduling, a +/- allowance of 50% for the 2 week and 6 week time points and 10% for 6, 12, 18 and 24 months was allotted. Fourteen patients received a cochlear implant or transferred out of the service within their first two years of life. In these cases, datalogging was collected up until the time point of their implantation or transfer.

### Sociodemographic information

Sex, ethnicity, and home language information was collected for each patient using Auditbase and Powerchart software (Oracle Cerner, USA) on the hospital system. Ethnicities were categorised as: Asian (including Asian Bangladeshi, Indian, Pakistani), Black (including Black Caribbean and African), Chinese, mixed ethnicity and white (including white British and other).

Home language was categorised as ‘English’ and ‘other language’. This was to allow comparison of families who use English as a primary language and families who do not. The hypothesis is that any language other than English may contribute to barriers for the family, rather than one specific language putting the family at higher risk than another language.

Therefore, sub-categorising language was not determined to be necessary at this stage.

Index of Multiple Deprivation Decile score (IMD), Income Decile score (ID), Education Decile score (ED) and Income Decile Affecting Children Index score (IDACI) was calculated using the patient’s home postcode. The score encompasses social factors such as income, education and health. Scores are numerical and rank from 1 (highest deprivation) to 10 (lowest deprivation).

### Statistical analysis

A statistical analysis was performed using the R 4.2.2 program package (Imperial College London, UK). Statistical significance was set to *p* =<0.05. The Shapiro-Wilk test found results to be non-normally distributed and therefore median, interquartile ranges and extreme values are presented. Non-parametric tests for statistical significance were employed.

Descriptive statistics were calculated from the datalogging values to explore average hearing aid use within the first two years post-fitting. Categorical clinical and socioeconomic variables were dichotomised due to the small sample size and analysed against average datalogging values using Wilcoxon Rank-Sum testing. Spearman’s correlation was performed to assess an association between continuous variables.

## Results

Ninety-four participants were recruited and datalogging measured at 6 different time points (2, 6 weeks, 6, 12, 18, 24 months post-fitting). There were 58 points missing due to patients who were either implanted (n=8), transferred out of the service post-fitting (n=8) or missed to follow up (n=8), resulting in a total of 506 potential data collection points. Datalogging was not consistently recorded during appointments, with 296 opportunities missed. This resulted in a total of 210 data points. From the records, it was not always obvious why datalogging had not been collected. Reasons noted by clinicians included patients not attending their appointment, hearing aids were lost and replaced or upgraded so datalogging information was not available. It is assumed that clinical time-pressure was the main barrier. A total of 32 participants had only one datalogging data point across the 6 time points. Twenty-seven had two; 19 had three; 13 had four and 3 participants had five. There were no participants with datalogging for all 6 time points.

Median age of initial fitting was 5.6 months (SD=5.77 months). The age of premature infants was recorded as corrected. The datalogging results indicated a median of 4.67 hours (3.0-7.3) per day use within the first two years post-fitting. There was insufficient data at this stage to determine how hearing aid use time changes within the first two years post-fitting.

Preliminary statistical analysis was performed to explore initial associations between datalogging and patients’ clinical and socioeconomic factors. They are outlined in table 1. It It was necessary to dichotomise the levels of the categorical variables at this phase. The mean “average datalogging” value used during analysis was calculated from the total datalogging values collected for each patient. SII scores for 15 participants fitted before 2013 were not viewable and the variable was therefore excluded during analysis. This was owing to restricted access to outdated software which carried these values.

**Table 1.** Comparison of datalogging values against categorical clinical and socio-economic factors (IMD=Index of Multiple Deprivation, IDACI= Income Decile Affecting Children Index)

| Variable | N/(%) | Median<br>datalogging<br>(hr/day) | Quartiles | Extremes | P-value |
| --- | --- | --- | --- | --- | --- |
| Clinical |  |  |  |  |  |
| *Uni vs bil* |  |  |  |  | 0.3 |
| Unilateral | 11 (12%) | 4.05 | 1.5-5.67 | 0-14 |  |
| Bilateral | 83 (88%) | 4.7 | 3.16-7.73 | 0.1-13.9 |  |
| *Additional disability* |  |  |  |  | 0.11 |
| No | 71 (76%) | 5 | 3.16-7.96 | 0-14 |  |
| Yes | 23 (24%) | 3.5 | 2.18-5.42 | 0.1-12.3 |  |
| Socio-economic |  |  |  |  |  |
| **Sex**<br>Female<br>Male | 46 (49%)<br>48 (51%) | 4.96<br>4.39 | 3.02-7.33<br>2.94-7.24 | 0.4-14<br>0-11.9 | 0.61 |
| **Home language**<br>Other language<br>English | 38 (40%)<br>56 (60%) | 4.1<br>5.25 | 2.89-6.36<br>3.23-7.96 | 0-10.5<br>0.2-14 | 0.3 |
| **Ethnicity**<br>BAME/other<br>White | 57 (61%)<br>37 (39%) | 4.05<br>5.62 | 3.03-6.8<br>3.5-7.5 | 0.1-14<br>0-13.9 | 0.17 |
| **IMD**<br>1-5<br>6-10 | 37 (39%)<br>57 (61%) | 4.31<br>5.5 | 2.23-5.98<br>3.25-8 | 0.1-13.9<br>0-14 | 0.19 |
| **Income decile**<br>1-5<br>6-10 | 46 (51%)<br>48 (49%) | 4.17<br>5.3 | 2.37-6.33<br>3.45-8 | 0.1-13.9<br>0-14 | 0.14 |
| **Education and skills decile**<br>1-5<br>6-10 | 23 (24%)<br>71 (76%) | 4.11<br>5.1 | 3.08-4.44<br>3.02-7.96 | 0.2-12.3<br>0-14 | 0.22 |
| **IDACI**<br>1-5<br>6-10 | 46 (49%)<br>48 (51%) | 4.05<br>5.85 | 2.0-8.53<br>3.5-8.2 | 0-12.3<br>0.4-14 | 0.01* |

\* Statistical significance $p$ value = $<0.05$

### Clinical variables

Median datalogging values for infants with unilateral and bilateral hearing aids were 4.05 (1.5-5.67) and 4.7 (3.16-7.73) hours per day respectively. For bilateral hearing aid users with differing datalogging values, the higher value was recorded. Those with disabilities, whether they were known at the time of fitting or not, showed a median of 3.5 (2.18-5.42) hours of use per day and infants with no known disabilities wore their hearing aids an average of 5 (3.16-7.96) hours. A Spearman’s coefficient of 0.17 showed a weak relationship between pure tone average thresholds and average datalogging values. All three variables did not reach statistical significance (*p=0*.*1-0*.*3*) and can be shown in table 1 and 2.

**Table 2.** Comparison of datalogging values against average pure tone threshold

| Variable | Coefficient | 95% CI | $P$-value |
| --- | --- | --- | --- |
| Average pure tone threshold | 0.17 | -0.03 to 0.36 | 0.1 |

### Socioeconomic factors

Four deprivation decile measurements were collapsed into two levels: 1 to 5, representing higher deprivation (lower opportunity) and 6 to 10, representing lesser deprived (higher opportunity). Infants whose families were placed in the higher deprivation group wore their hearing aids for 4.31 (2.23-5.98), 4.17 (2.37-6.33), 4.11 (3.08-4.44), 4.05 (2.0-8.53) hours per day according to the Index of Multiple Deprivation Decile, Income Decile, Education and Skills Decile and Income Deprivation Affecting Children (IDACI) respectively. Those who fell into the lower deprivation group achieved 5.5 (3.25-8), 5.3 (3.45-8), 5.1 (3.02-7.96), 5.85 (3.5-8.2) hours per day for the same. A statistically significant difference between infants in the higher and lower deprivation groups according to IDACI was observed (*p=0*.*01*). There was no concern of familywise error as significance reached a p-value of 0.01. No other differences reached significance (*p=0*.*14-0*.*22)*.

By ethnicity, infants from white backgrounds, including white British and white other achieved 5.62 (3.5-7.5) hours per day use, while those from Black, Asian and minority ethnic and other ethnicity groups wore their hearing aids for 4.05 (3.03-6.8) hours per day. By home language, infants whose families used English as their primary language wore their hearing aids for 5.25 (3.23-7.96) hours per day and those from families who use another language other than English achieved 4.1 (2.89-6.36) hours.

The smallest difference in datalogging values between variables was observed according to sex, with female and male infants achieving 4.96 (3.02-7.33) and 4.39 (2.94-7.24) average hours per day use respectively. Differences in datalogging values according to ethnicity, home language and sex were not found to be statistically significant (*p=0*.*17-0*.*61*).

**Figure 1.**
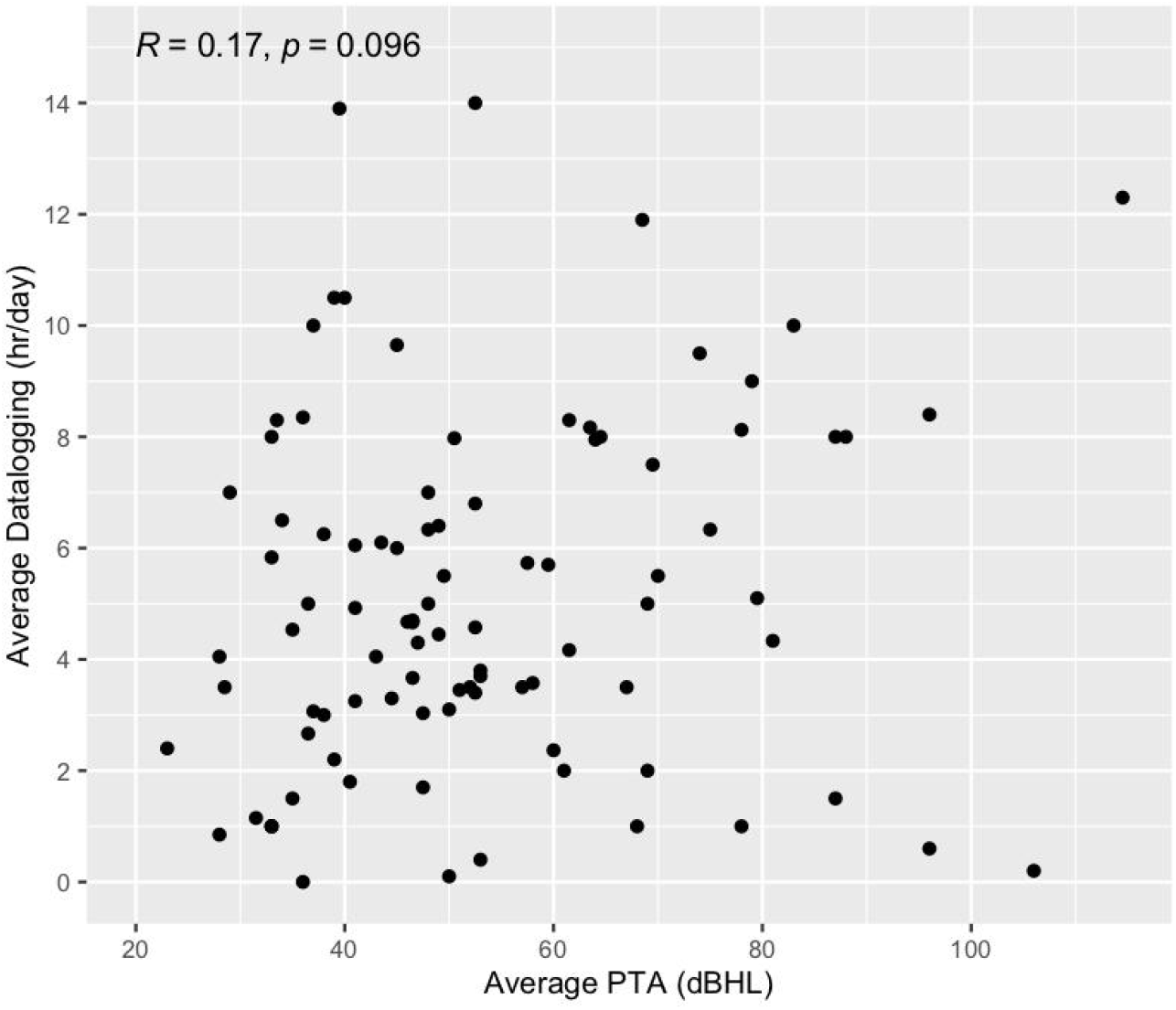
Relationship between 3 or 4 frequency average pure tone thresholds and daily average datalogging

**Figure 2.**
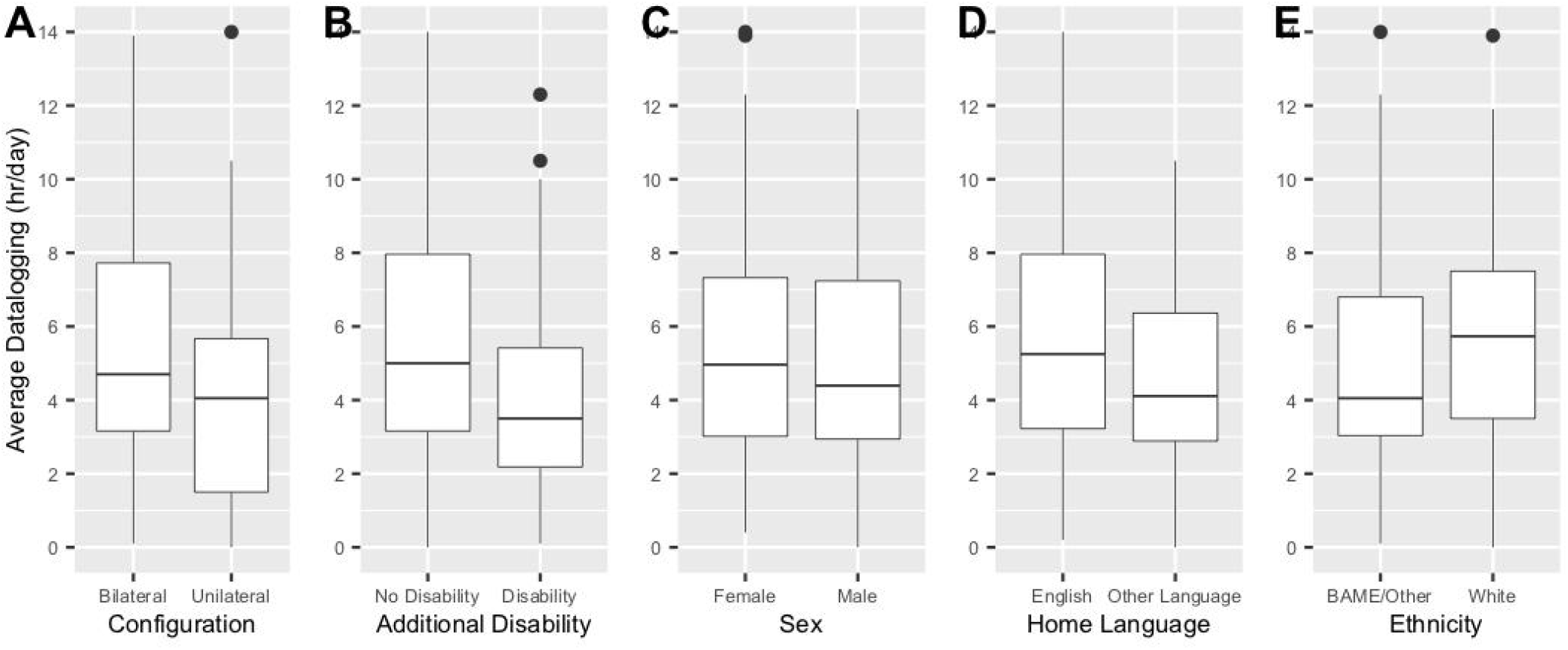
Datalogging values by categorical clinical and sociodemographic variable: (A) lateral configuration, (B) additional disability, (C) sex, (D) home language, (E) ethnicity. The box bounds the interquartile range (IQR) divided by the median. Tukey-style whiskers extend to a maximum of 1.5x IQR with filled circles representing data >1.5x IQR.

**Figure 3.**
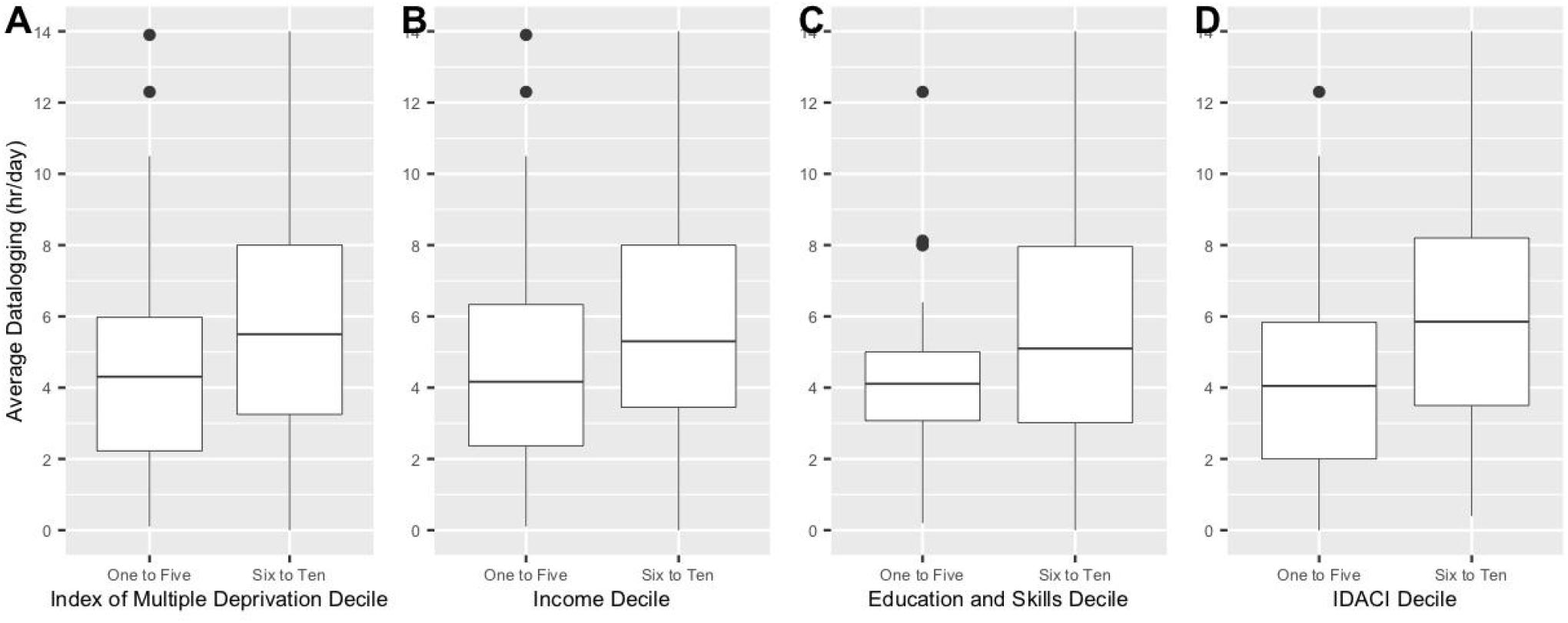
Datalogging values by Deprivation Decile: (A) Index of Multiple Deprivation (IMD) (B) Income Decile (C) Education and Skills Decile (D) Income Decile Affecting Children Decile (IDACI)

## Discussion

Findings show that infants in our cohort are achieving less than optimal usage of their hearing aids, putting them at risk of the later effects associated with limited exposure to speech and language during what is a critical period in their development. Our results are in line with existing literature which show similar values when datalogging is explored [6,12].

Although most of the results were under-powered to reach statistical significance, there is some preliminary evidence to suggest that infants whose families may experience barriers to healthcare provision due to language, ethnicity and socioeconomic status may achieve less than optimal usage of their hearing aids by up to 108 minutes per day within the first two years of life. Those who have additional disabilities or use a unilateral hearing aid also show lower use.

Our findings have wider implications on practice and suggest that there are not only clinical and social factors involved with optimal usage of hearing aids, but that the individual needs of infants and their families may not be addressed. As healthcare moves into a culture of ‘individualised’ and ‘family-centred’ care, more work is needed to understand and address the complex and diverse elements of truly equitable paediatric audiology services.

### Datalogging use in clinic

In this single-site pilot study, datalogging values were not consistently collected at the available time points in clinic. This is consistent with previous reports of 43% of audiologists always checking datalogging at follow-up [20]. However, this is based on datalogging use in adult clinics, with use in paediatric clinics is not well understood. Reasons for inconsistent collection of datalogging noted by the clinicians included lost hearing aids where datalogging information was not accessible and unattended appointments. Time constraints within appointments are thought to be the primary barrier. Variation in datalogging collection by audiologists can be explored in a larger scale study to better understand its utilisation and associated barriers. This study flags a possible need for additional training of audiologists to promote and prioritise datalogging as a counselling tool, but strategies to address cultural, social and clinical barriers to usage with families are still poorly understood. In audiology, we are the envy of many other healthcare specialisms in that we have an objective, quantitative measure of adherence to treatment and their value to research should also not be overlooked.

### Limitations

Clinicians at this centre are from diverse backgrounds and are native speakers of additional languages. Some appointments could have been conducted in the parents’ preferred language whereas use of interpreters, although widespread, may not have been available for some families of a linguistic minority. A larger scale study incorporating rural services and those with a more homogenous population could yield larger differences in hearing aid use where home language is concerned. Fluency of English was also not measured in families who use an additional language, meaning that diversity within the variable is not well considered.

Using decile scores from the Index of Multiple Deprivation based on 2019 data is not an exhaustive measure of socioeconomic status and comes with limitations. In urban London where the study was based, ‘lower layer super output areas’ or LSOAs are subject to rapid change and gentrification is not necessarily captured. It also fails to address the intersection of ethnicity, socioeconomic status, education and income to the individual. It is however a useful tool used widely in research and service strategy but should be interpreted with caution.

### Future research

While valuable, the findings from this study are based on incomplete data from a single site. A larger-scale study incorporating multiple sites from a variety of geographical locations is needed to further explore the impact of clinical and sociodemographic factors of hearing aid use in infants within the first two years. Our findings show that hearing aid use in our cohort is less than optimal, putting infants at risk of the later effects associated with limited exposure to speech and language during what is a critical period in their development. High quality research is needed to understand the barriers and facilitators to hearing aid use in families of all backgrounds, to capture their needs and develop services to deliver equitable habilitation plans and support.

## Conclusions

Average datalogging values for daily hearing device use were lower than considered optimal for 0-2 year olds with permanent childhood deafness. The only factor related to these findings was socioeconomic status indicating that infants from more deprived backgrounds may achieve lower hearing aid usage than those from the lesser deprived regions. This finding needs to be verified on a larger scale and better understood to explore potential approaches to overcome the problems. The role of multiple factors also needs to be explored with a larger sample size. This would require a multi-centre study to be conducted.

## Data Availability

All data produced in the present study are available upon reasonable request to the authors.

## Funding

No external funding was directly provided for this study. EG is undertaking a funded HEE/NIHR Predoctoral Clinical Academic Fellowship (NIHR302719). D.A.V is funded by an MRC Senior Fellowship in Hearing (MR/S002537/1) and NIHR programme grant for applied research (201608).

## Notes

### Competing Interest Statement

The authors have declared no competing interest.

### Funding Statement

No external funding was directly provided for this study. E.G is undertaking a funded HEE/NIHR Predoctoral Clinical Academic Fellowship (NIHR302719). D.A.V is funded by an MRC Senior Fellowship in Hearing (MR/S002537/1) and NIHR programme grant for applied research (201608).

### Author Declarations

Health Research Authority and Health and Care Research Wales gave ethnical approval for this work.

## References

1. Wood, S. A., Sutton, G. J., & Davis, A. C. (2015). Performance and characteristics of the newborn hearing screening programme in England: The first seven years. International Journal of Audiology, 54(6), 353–358. https://doi.org/10.3109/14992027.2014.989548

2. Fortnum, H., & Davis, A. (1997). Epidemiology of Permanent Childhood Hearing Impairment in trent region, 1985–1993. British Journal of Audiology, 31(6), 409–446. https://doi.org/10.3109/03005364000000037

3. Ching, T. Y. C., Dillon, H., Button, L., Seeto, M., Van Buynder, P., Marnane, V., Cupples, L., & Leigh, G. (2017). Age at intervention for permanent hearing loss and 5-year language outcomes. Pediatrics, 140(3). https://doi.org/10.1542/peds.2016-4274

4. Moeller, M. P. (2007). Current state of knowledge: Psychosocial development in children with hearing impairment. Ear & Hearing, 28(6), 729–739. https://doi.org/10.1097/aud.0b013e318157f033

5. Walker, E. A., McCreery, R. W., Spratford, M., Oleson, J. J., Van Buren, J., Bentler, R., Roush, P., & Moeller, M. P. (2015). Trends and predictors of longitudinal hearing aid use for children who are hard of hearing. Ear & Hearing, 36(Supplement 1). https://doi.org/10.1097/aud.0000000000000208

6. Jones, C., & Launer, S. (2010). Pediatric fittings in 2010: The sound foundations cuper project. A sound foundation through early amplification, 1020.

7. Fitzpatrick, E. M., Durieux-Smith, A., & Whittingham, J. (2010). Clinical practice for children with mild bilateral and unilateral hearing loss. Ear & Hearing, 31(3), 392–400. https://doi.org/10.1097/aud.0b013e3181cdb2b9

8. Moeller, M. P., Hoover, B., Peterson, B., & Stelmachowicz, P. (2009). Consistency of hearing aid use in infants with early-identified hearing loss. American Journal of Audiology, 18(1), 14–23. https://doi.org/10.1044/1059-0889(2008/08-0010)

9. Muñoz, K., Blaiser, K., & Barwick, K. (2013). Parent hearing aid experiences in the United States. Journal of the American Academy of Audiology, 24(01), 005–016. https://doi.org/10.3766/jaaa.24.1.2

10. Muñoz, K., Roberts, M., Mullings, D., & Harward, R. (2012). Parent hearing aid experience. The Volta Review, 112(1), 63–76.

11. Marnane, V., & Ching, T. Y. C. (2015). Hearing aid and cochlear implant use in children with hearing loss at three years of age: Predictors of use and predictors of changes in use. International Journal of Audiology, 54(8), 544–551. https://doi.org/10.3109/14992027.2015.1017660

12. Walker, E. A., Spratford, M., Moeller, M. P., Oleson, J., Ou, H., Roush, P., & Jacobs, S. (2013). Predictors of hearing aid use time in children with mild-to-severe hearing loss. Language, Speech, and Hearing Services in Schools, 44(1), 73–88. https://doi.org/10.1044/0161-1461(2012/12-0005)

13. Muñoz, K., Olson, W. A., Twohig, M. P., Preston, E., Blaiser, K., & White, K. R. (2015). Pediatric hearing aid use: parent reported challenges. Ear & Hearing, 36(2), 279–287. https://doi.org/10.1097/aud.0000000000000111

14. Visram, A. S., Roughley, A. J., Hudson, C. L., Purdy, S. C., & Munro, K. J. (2020). Longitudinal changes in hearing aid use and hearing aid management challenges in infants. Ear & Hearing, 42(4), 961–972. https://doi.org/10.1097/aud.0000000000000986

15. Caballero, A., Muñoz, K., White, K., Nelson, L., Domenech-Rodriguez, M., & Twohig, M. (2017). Pediatric hearing aid management: Challenges among Hispanic families. Journal of the American Academy of Audiology, 28(08), 718–730. https://doi.org/10.3766/jaaa.16079

16. Kubba, H., Macandie, C., Ritchie, K., & Macfarlane, M. (2004). Is deafness a disease of poverty? the association between socio-economic deprivation and congenital hearing impairment. International Journal of Audiology, 43(3), 123–125. https://doi.org/10.1080/14992020400050017

17. Sutton, G. J., & Rowe, S. J. (1997). Risk factors for childhood sensorineural hearing loss in the oxford region. British Journal of Audiology, 31(1), 39–54. https://doi.org/10.3109/03005364000000007

18. Czechowicz, J. A., Messner, A. H., Alarcon‐Matutti, E., Alarcon, J., Quinones‐Calderon, G., Montano, S., & Zunt, J. R. (2010). Hearing impairment and poverty: The epidemiology of ear disease in Peruvian schoolchildren. Otolaryngology–Head and Neck Surgery, 142(2), 272–277. https://doi.org/10.1016/j.otohns.2009.10.040

19. Almazroua, A. M., Alsughayer, L., Ababtain, R., Al-shawi, Y., & Hagr, A. A. (2020). The association between consanguineous marriage and offspring with congenital hearing loss. Annals of Saudi Medicine, 40(6), 456–461. https://doi.org/10.5144/0256-4947.2020.456

20. McMillan, A., Durai, M., & Searchfield, G. D. (2017). A survey and clinical evaluation of Hearing Aid Data-logging: A valued but underutilized hearing aid fitting tool. Speech, Language and Hearing, 21(3), 162–171. https://doi.org/10.1080/2050571x.2017.1339942

